# Report: Evaluating the ability of the TMA Clinical Decision Support platform to identify interventions in a clinical setting

**DOI:** 10.1101/2021.05.24.21257617

**Authors:** Joshua Resnikoff, Yessica Giraldo, Lina Williamson

## Abstract

The TMA Precision Health Clinical Decision Support system is a commercially available software platform focused on the confirmation of a precision diagnosis and generation of a personalized care plan to rapidly deliver therapeutic optionality and improve quality of life for rare and complex disease patients. For this study, we worked with our partners in Medellin, Colombia to evaluate the efficacy of the platform in identifying previously unexplored modes of care within a small sample population of adult patients suffering from a diverse set of rare diseases. Although challenges were encountered during the curation of data from multiple sources, personalized care plans and medication options were identified successfully for 94% of cases, suggesting a high level of impact for deployment at scale.

## Background

Individual rare diseases may affect only a limited number of patients, but rare disease patients as a group are estimated to account for up to as much as 8% of the general population [1]. For most medical practitioners, rare or orphan disease patients represent the most difficult to diagnose and treat cases, and therefore the most complicated to manage [2]. Physicians have limited training throughout the course of their career to prepare them for the challenges associated with diagnosing and managing rare diseases [3], which can exacerbate poor decision making and ineffective outcome-driven communication during the patient-physician encounter [4]. General practitioners (GP) self-report both the lowest awareness of rare disease knowledge and a majority lack awareness about investigational resources like Orphanet [5]. But even access to basic knowledge is not enough: it has been reported that GPs lack a comprehensive or systematic primary care approach to treat rare diseases [6], and in fact do not physically have the time to meet current care guideline recommendations for well-defined diseases, even when disease progression was controlled for “stable” and “maintenance phase” [7]. Taken together, the challenges related to information access for rare diseases are the largest impediment to effective rare disease management [8], in turn exacerbating disease progression [9], misdiagnosis, and patient morbidity [10].

Providing Clinical Decision Support (CDS) tools to physicians has demonstrated improvement in the efficiency, delivery, and quality of care [11]. The evidence generated from CDS systems employed in oncology management strongly suggests that the practice of personalized precision healthcare improves outcomes for patients [12]. To our knowledge, there is no relevant preceding technology to act as a CDS system for GPs encountering rare disease patients during their career. As a result, the rare disease patient experience is among the least efficient – requiring an average of 7.3 physicians and 4.8 years to achieve diagnosis [13] - and most costly – accounting for nearly $1 trillion in annual costs in the US alone [14].

TMA Precision Health offers an AI-based platform to provide rare disease patients and their GP personalized and actionable therapeutic and care management options to improve patients’ and their families’ quality of life more rapidly than the current paradigm. In this study we conducted a preliminary deployment of the technology into a random sample of adult Colombian rare disease patients to evaluate the platform’s performance and capacity to deliver CDS plans to this community.

## Methods

### Case identification

We partnered with one of the largest health insurers in Colombia to identify rare disease patients that represented high-cost individuals due to challenges in disease understanding and/or mismanagement. The insurer supplied a list of their top 100 non-pediatric high-cost patients without a genetic diagnosis that also utilized Promedan IPS as a primary source of care. 28 cases were selected based on the completeness of electronic medical histories. 7 cases were randomly selected and held back as potential alternates, with the remaining 21 contacted via the Promedan Data Manager to receive consent to participate in the study. 10 participants were included in the final study as a function of consent and our ability to schedule interviews with them to better understand the scope of their medical history.

### Patient consent

To comply with Colombian regulations regarding patient privacy, TMA created a Colombian based joint venture with Promedan IPS, a private clinic in Medellin, Antioquia, called ION Health. Patients were contacted and consented in coordination with Colombian regulations administered through the professional staff at Promedan IPS.

### IRB Approval

Study design and patient consent was approved by an independent review board of the Universidad CES in Medellin, Antioquia.

### Diagnosis and medication review

Anonymized patient medical history was captured in the ION Health electronic heath repository and manually curated to remove duplicate entries arising from multiple originating sources within the Colombian healthcare system (hospitals, labs, insurers, etc.). Available lifetime verified medical and clinical diagnoses were tallied on a per case basis. TMA Case Coordinators then used the TMA platform to review the patient case medical history and leading diagnosis against published disease descriptions accessed through the TMA platform (powered by LifeMap Sciences, Yeda Research and Development Company, Ltd, Israel). Case specific medications were organized into a Gantt format and then tallied on a per case basis.

### Identification of therapeutic alternatives

Using the leading diagnosis of each case as the basis for further investigation, we utilized our platform NLP to label the patient case history based on the standard medical ontologies of ICD-10-CM and RXNorm. Labels were then used to power predicate-driven search paths that output cause-and-effect relationships between medication, genetics, and phenotype, which can be cross-referenced against the patient history. Previously unexplored and/or unreported care plans and/or medications were captured for inclusion in the final report. Suggested care protocols and/or medications were reviewed internally by a panel of medical doctors to verify result accuracy prior to publication and dissemination to the patient’s GP.

## Results

Colombia is home to an estimated 49,000,000 people, comprising a rare disease patient population of approximately 3,920,000 patients (8% global estimate). For this study, we reviewed a sample of adult patients with a likely confirmed medical diagnosis (non-genomic based) identified by their insurance provider and receiving care from our clinical partner, to evaluate the use of our platform for the purposes of documenting previously unidentified care management and medication options that have been demonstrated to improve the quality of life and/or disease outcomes in corresponding patient phenotypes. Our sample population (n=10, average age 58) suffers from a wide array of symptoms (average = 31) and disease states (average = 14) that have been under review for years to decades (average = 20), while attempting to gain some level of permanent relief from a wide array of medications (average = 22)(Figure 1).

**Figure 1.**
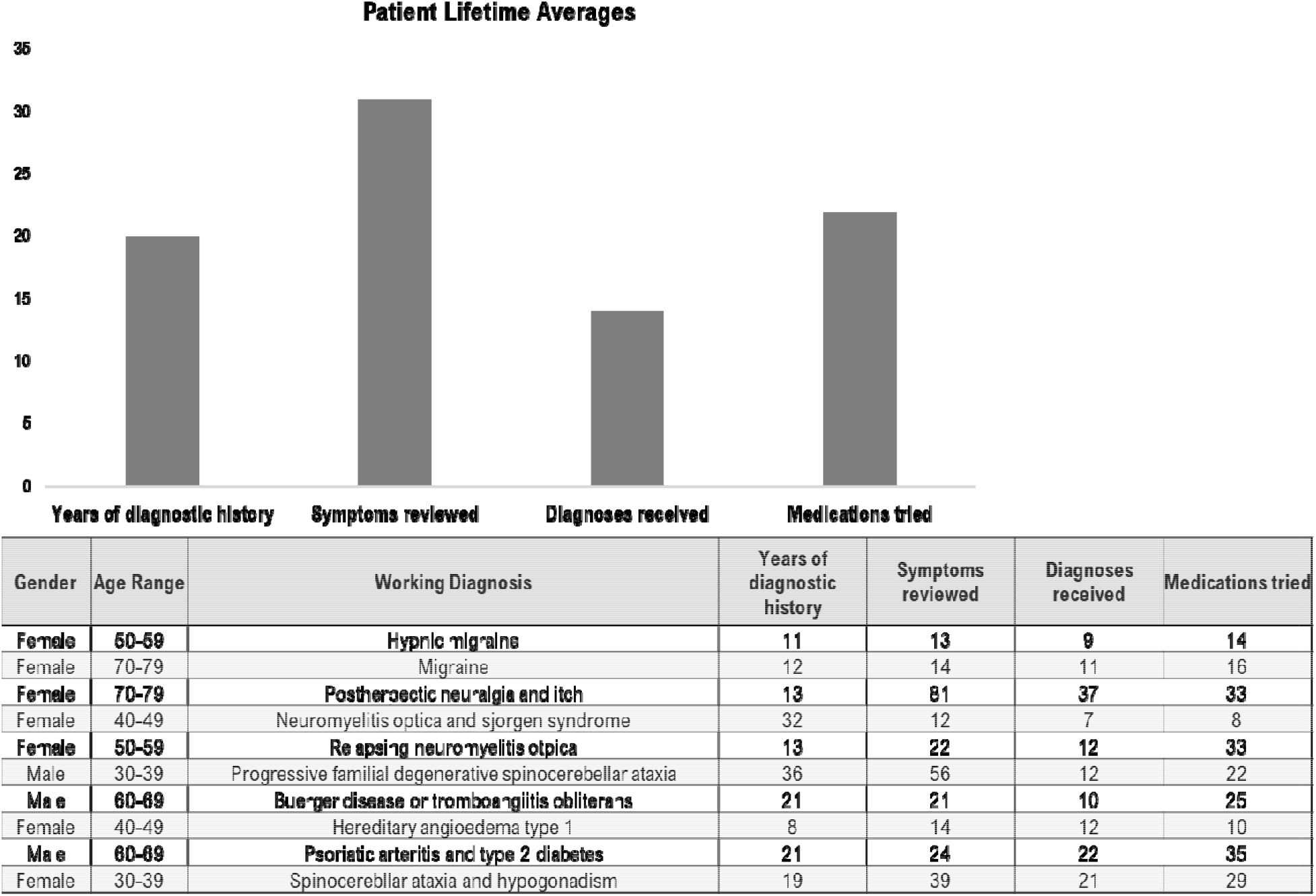
De-identified informal on detailing the individual and average lifetime available histories for the health care medical odyssey endired by rare disease patients in the sample population, n=10. Average age of study participant is 58; average years experienced searching for a diagnoses is 20; average symptoms reported over that timeframe is 31; average alternate diagnoses received over that time period is 14; the average medications tried responding to these various diagnoses is 22.

Individual cases were reviewed and personalized precision case reports comprising care protocols and previously untried medications were generated. Once a patient’s data had been entered into the Ion EHR and curated, it took a TMA Case Coordinator an average of 12 hours to review and process the information to create a personalized precision intervention report. Across 100% of cases, the TMA platform was able to identify preventive and/or treatment care plans that had not been utilized by the patient previously. In 90% of cases (9 of 10 cases, n=10), the TMA platform was able to identify medications – including repurposed and disease-specific – that have been shown to positively effect health outcomes in patients suffering from the same disease (Table 1). Taken together this represents a 95% success rate in TMA’s ability to rapidly identify previously untried interventions, despite multiple decades worth of doctors’ time and resources committed to these patientos within an advanced clinical setting.

**Table 1:** Outcomes related to the hours required to identify care protocals and medicines that have citable positive impact on the health outcomes of patients with matched phenotypes.

| Gender | Age Range | Working Diagnosis | TMA Care Protocols | Alternate medication options | Case hours to complete |
| --- | --- | --- | --- | --- | --- |
| Female | 50-59 | Hypnic migraine | 2 | 3 | 6 |
| Female | 70-79 | Migraine | 1 | 6 | 8 |
| Female | 70-79 | Postherpetic neuralgia and itch | 6 | 13 | 24 |
| Female | 40-49 | Neuromyelitis optica and sjorgen syndrome | 5 | 7 | 6 |
| Female | 50-59 | Relapsing neuromyelitis optica | 3 | 5 | 6 |
| Male | 30-39 | Progressive familial degenerative spinocerebellar ataxia | 10 | 7 | 40 |
| Male | 60-69 | Buerger disease or thromboangiitis obliterans | 3 | 4 | 8 |
| Female | 40-49 | Hereditary angioedema type 1 | 3 | 9 | 8 |
| Male | 60-69 | Psoriatic arthritis and type 2 diabetes | 9 | 0 | 8 |
| Female | 30-39 | Spinocerebellar ataxia and hypogonadism | 3 | 1 | 8 |

## Discussion

For a patient or patient family encountering a life-changing illness it is paramount they have the confidence that their GP is considering all the relevant and cutting-edge data to make diagnostic and therapeutic decisions in a timely manner [3], but this is difficult to achieve in all but the most unique instances of care, perpetuating the challenges rare disease patients experience which manifests in direct and indirect costs on the US health system of nearly $1 trillion dollars [14]. While there continues to be a growing interest in developing boutique solutions for specific rare diseases, these efforts are constrained primarily at only the largest and most well-funded academic institutions, leaving millions of patients without direct access to disease specialists who have knowledge of their specific condition [15] and their GPs without access to the self-reported resources needed to rapidly understand and develop a care plan for a rare disease [6, 7].

The largest challenge encountered within this study was the lack of interoperability and need for hands-on curation of patient data to formulate a succinct and comprehensive patient medical history. While challenges will persist around the intake and curation of data from unorganized originating sources in geographies that lack access to, or have not yet implemented, digitally accessible infrastructure, we anticipate this issue will not present as significantly in countries that have had the opportunity to incorporate robust electronic medical record systems into their standard workflows. Moreover, understanding this specific challenge allows us to integrate solutions that supersede this digital divide and address interoperability from the outset, via either the custom development or software partnerships to gather and curate patient medical information according to our preferred data structure.

Our study demonstrates the feasibility of utilizing the TMA platform in a clinical setting to develop clinical decision support strategies that can provide immediate benefit to both the GP and the rare disease patient. The wide array of diseases that we encountered within such a small sample size is indicative of the diversity of the 7,000+ rare diseases that affect this grouped patient cohort. The high determined rate of 94% intervention and care identification implies the breadth of unexplored knowledge that lies beyond the immediate grasp of GPs, possibly because of their existing work burden, their inexperience in encountering rare disease(s), and/or their lack of a systemic approach to creating a care plan to effectively manage these types of patients. Understanding the need for novel solutions to meet the anticipated EHR gap in ex-US locations underscores the customizable instantiation of the system, which will improve the ability of the platform to support globally diverse GPs. All of this suggests that the TMA platform and process is scalable across a multitude of rare disease states and geographic locations, for the potential betterment of the patient health and the increased effectiveness of front-line physicians. Continued exploration identifying and transporting standards-of-care across international lines will only serve to further the potential impact on the quality of life and healthcare experience outcomes for rare disease patients around the world.

## Data Availability

The authors confirm that the data supporting the findings of this study are available within the article.

## Notes

Sponsor: Ion Health, the Precision Medicine Institute of Latin America, Medellin Antioquia Colombia

### Competing Interest Statement

TMA Precision Health is a partner of ION Health and helped underwrite this study.

### Funding Statement

The author(s) received no financial support for the research, authorship, and/or publication of this article.

### Author Declarations

IRB approval was received by an independent IRB Review Board at the Universidad CES in Medellin, Antioquia, Colombia.

